# A cost-effectiveness of Fecal DNA methylation test for colorectal cancer screening in Saudi Arabia

**DOI:** 10.1101/2022.11.15.22282325

**Authors:** Zhongzhou Yang, Mang Shi, Mengping Liu, Zhe Wang, Hui Huang, Shunyao Wang, Xiaoyuan Zheng, Yanyan Liu, Na Liu, Yantao Li, Eric Lau, Shida Zhu

## Abstract

**Background:** In the Saudi Arabia, we estimated the cost-effectiveness between fecal DNA methylation test (FDMT) and fecal immunochemical testing (FIT) to detect colorectal cancer (CRC) and precancerous lesions in the national screening program.

**Participants and methods:** A Markov model was used from 45 to 74 years old CRC screening to compare the cost-effectiveness with the FDMT vs FIT. We predicated the longitudinal participation patterns in the perfect adherence vs organized programs screening covered by national budgets. Outcomes incorporated the incidence rates and mortality rates, cost, quality-adjusted life-years (QALYs), incremental cost-effectiveness ratios (ICERs) under the perfect adherence as well as incidence and mortality forecast within 3, 6 and 9 years.

**Results:** Under the perfect adherence, the total cost of FDMT was cheaper 38.16% than FIT and extends 0.22 QALYs per person. Furthermore, FDMT was more cost-effective as ICERs ($1487.30 vs $1982.42 per QALY saved) compared with FIT test. Therefore, FDMT test dominated than FIT every year (more costly and less effective). Compared with the organized FDMT programs (6.6% initial positive rate and 54% coloscopy compliance rate), the FIT program (5.8% initial positive rate and 48% coloscopy compliance rate) had 6.25 times to 7.76 times on the incidence rates; 5.12 times to 12.19 times on the mortality rates among 3, 6 and 9 years prediction.

**Conclusions:** Through the Markov model, we compared FDMT was less costly and more effective than the FIT test under the perfect and organized adherence within nine years prediction. It implied that FDMT might the novel cost-effective tool for Saudi Arabia national screening program.

## Introduction

According to WHO, Colorectal cancer (CRC) is the third most common cancer in 2020 and led to 9.0 per 100, 000 mortality rate from age 50 to 74 [1-3]. Specifically, CRC was the largest death number as 1996 among all cancers in Saudi Arabia [4]. The large-scale clinical trials have shown the effectiveness of fecal immunochemical testing (FIT) to reduce the burden of CRC. However, it was not sufficient because of three factors. The first reason was short of the evidences to support for FIT in the national screening, though the small 9% proportion of citizens experienced screening based on their healthcare providers or their own interests in Saudi Arabia [5, 6]. Secondly, general population lack of the awareness about the common risk factors and the potential benefits of CRC screening from doctor recommendations in Saudi Arabia [7, 8]. Thirdly, the priority strategy was determined by three factors incorporating the national budget, colonoscopy capacity and cost of per QALY [9]. Hence, it is necessary to develop the new screening test especially for the national program.

### DNA methylation testing

Fecal DNA methylation test (FDMT) is developed as the novel fecal test (ColoTect 1.0 version; Beijing Genomics Institute) to screen colorectal cancer, and the large-scale clinical trials were undergoing in the national level recommended triennially as a complementary for FIT. FDMT had the additional benefits to detect the early and terminal stages of CRC. Meanwhile, FDMT is self-sampling without medication/diet restrictions and can be conducted at home. The previous project measured performance like sensitivity of FDMT in Qingdao population from China. The first FDMT version claimed sensitivity for precancerous lesion (46.0% vs 24 %), early stages (87.7% vs 70 %) and terminal stages (88.4% vs 76.5%) compared with FIT screening based on the provincial screening program in a total of 18,136 subjects at screening ages of 45 to 74.

This study aims to explore the screening cost effectiveness for precancerous lesions of colorectal cancer, early stages and terminal stages with FDMT compared with the current FIT screening strategy. Multiple Markov model have compared the health economic impact of the CRC screening modes. However, the previous studies seldom considered the complicated pattern of screening adherence and coloscopy compliance, which were more practical for the organized screening than the perfect screening pattern. Therefore, we conducted a Markov health economic model covering the assumed perfect participation or the complicated participation patterns of adherence and coloscopy compliance sourced from the real world, allowing for the prediction of colorectal cancer incidence rates and mortality rates within 9 years and incorporating the extra early and terminal stages predicated above.

## Methods

### Study design

We firstly applied the Markov model to simulate CRC screening in the standard 100,000 Saudi Arabia population to compare the triennially Fecal DNA methylation test and annual FIT screening patterns in nine years. The test performance considered different characteristics from age 45 to 75, and the predicated models for perfect adherence (100%) vs the organized screening (5.8%) from our previous project. First, we head to head compared the cost effectiveness of triennially Fecal DNA methylation test and annual FIT to determine the ICER in Saudi Arabia, under the assumption of perfect (100%) adherence. Second, we assumed five categories as normal group, 7% precancerous lesion group, 45 per 100,000 early-stage colorectal cancer (ES-CRC) of stages I and II, 40 per 100,000 terminal stage (TS-CRC) colorectal cancer of stages III and IV, and death group. Third, we simulated the number of CRC cases and deaths within nine years under the organized screening. Fourth, we estimated the indirect cost for national screening covering publicity, training, community action and screening labor fees. Our model consisted of the four major components: test performance, clinical component, cost component and health state utility component.

### Markov model and Screening

We established one Markov simulation model of the transition matrix (Appendix) succeeding the previous early screening models [10]. Our model optimized the assumptions with triennial screening pattern for FDMT. The screening pattern was followed by MT-sDNA from 3 CISNET models [11] as well as comparing from every five years to every year [12]. The test performance was claimed in Table 1 for Fecal DNA methylation test from research team including the novel sensitivity for 46% precancerous lesion, 87.7% stages I and II, 88.4% stages III and IV. FIT sensitivity was 24% precancerous lesion, 70% stages I and II, 74% ∼ 79% stages III and IV based on literature [13]. Health state utility were categorized by stages from one of normal status, 0.85 of precancerous lesions, 0.74 of stage I, 0.67 of stage II, 0.5 of stage III and 0.25 of stage IV.

**Table 1.** Parameters for the model input in Saudi Arabia 2020

| Table 1. Parameters for the model input in Saudi Arabia 2020 |  |  |
| --- | --- | --- |
| Variable | Value | References |
| Test performance |  |  |
| Fecal DNA methylation test sensitivity for precancerous lesion | 46% | Claimed |
| Fecal DNA methylation test sensitivity for stages I + II | 87.7% | Claimed |
| Fecal DNA methylation test sensitivity for stages III + IV | 88.4% | Claimed |
| Fecal DNA methylation test screening adherence rate | 5.8% | Qingdao |
| Fecal DNA methylation test participation compliance rates<sup>1</sup> | 47% | Qingdao |
| FIT sensitivity for precancerous lesion | 24% | Florence et al |
| FIT sensitivity for stages I + II | 70% | Product website |
| FIT sensitivity for stages III + IV | 74%~79% | Florence et al |
| FIT screening adherence rate | 6.6% | Saudi Arabia |
| FIT participation compliance rates<sup>1</sup> | 20% | Saudi Arabia |
| Clinical |  |  |
| Incidence for precancerous lesions | 7% | Article |
| Incidence for stages I + II | 45 per 100,000 | WHO & article |
| Incidence for stages III + IV | 40 per 100,000 | WHO & article |
| Annual transition rate to precancerous from normal | 1.50% | Uri |
| Annual transition rate to stages I + II from precancerous | 5% | Uri |
| Annual transition rate to stages III + IV from stages I + II | 8.30% | Uri |
| Mean survival from stages I + II | 4.5 y | Uri |
| Mean survival from stages III + IV | 2.8 y | Uri |
| Age to start screening | 45 | Uri |
| Age to stop screening | 74 | Uri |
| Cost |  |  |
| Fecal DNA methylation test direct cost | 102.3 | Estimated |
| Fecal DNA methylation test indirect (program) | 51 | Estimated |
| FIT direct | 27 | Government |
| FIT indirect cost (program) | 51 | Estimated |
| Coloscopy with Anesthesia | 2900 | Steffie-SJG-2021 |
| Stage 1 and 2 | 24,829 | Steffie-SJG-2021 |
| Stage 3 and 4 | 326,237 | Steffie-SJG-2021 |
| Health state utilities |  |  |
| Normal status | 1 | Dietz et al., 2003 |
| Precancerous lesions | 0.85 | Dietz et al., 2003 |
| Stage I | 0.74 | Dietz et al., 2003 |
| Stage II | 0.67 | Dietz et al., 2003 |
| Stage III | 0.5 | Dietz et al., 2003 |
| Stage IV | 0.25 | Dietz et al., 2003 |
| <sup>1</sup> Coloscopy after Fecal DNA methylation test or FIT positive |  |  |

### Cost-Effectiveness Analyses

We established the represented 100,000 virtual SA population regarding age, screening outcome, CRC incidence and mortality within nine years. The analysis was conducted in R (4.0.4) and we calculated the total cost, QALYs/person, cost/person incremental cost-effectiveness ratios (ICER) for the assumed perfect adherence. To conduct the cost-effectiveness analyses for the organized screening, we predict the probability, numbers and cost of FDMT and FIT screening for the outcome of normal, PL-CRC and CRC. Furthermore, both incidence rate and mortality rate were calculated per 100, 000 for three, six and nine years comparing between FDMT and FIT screening.

## Results

### Disease progression

For the disease progression, they included 7 compartments from normal status to death status in Figure 1. It had five categories as normal group, precancerous lesion group, early-stage colorectal cancer of stages I and II, terminal stage colorectal cancer of stages III and IV, and death group. The transition rate was 1.5% (*P*_*1*_) from normal group to precancerous lesion in annually [12, 14, 15]. Then, the annual transition rate was 5% (*P*_*2*_) from precancerous lesions to early-stage colorectal cancer [16, 17]. Furthermore, the annual transition rate was 8.3% (*P*_*3*_) from early-stage colorectal cancer to terminal-stage colorectal cancer.

**Figure 1.**
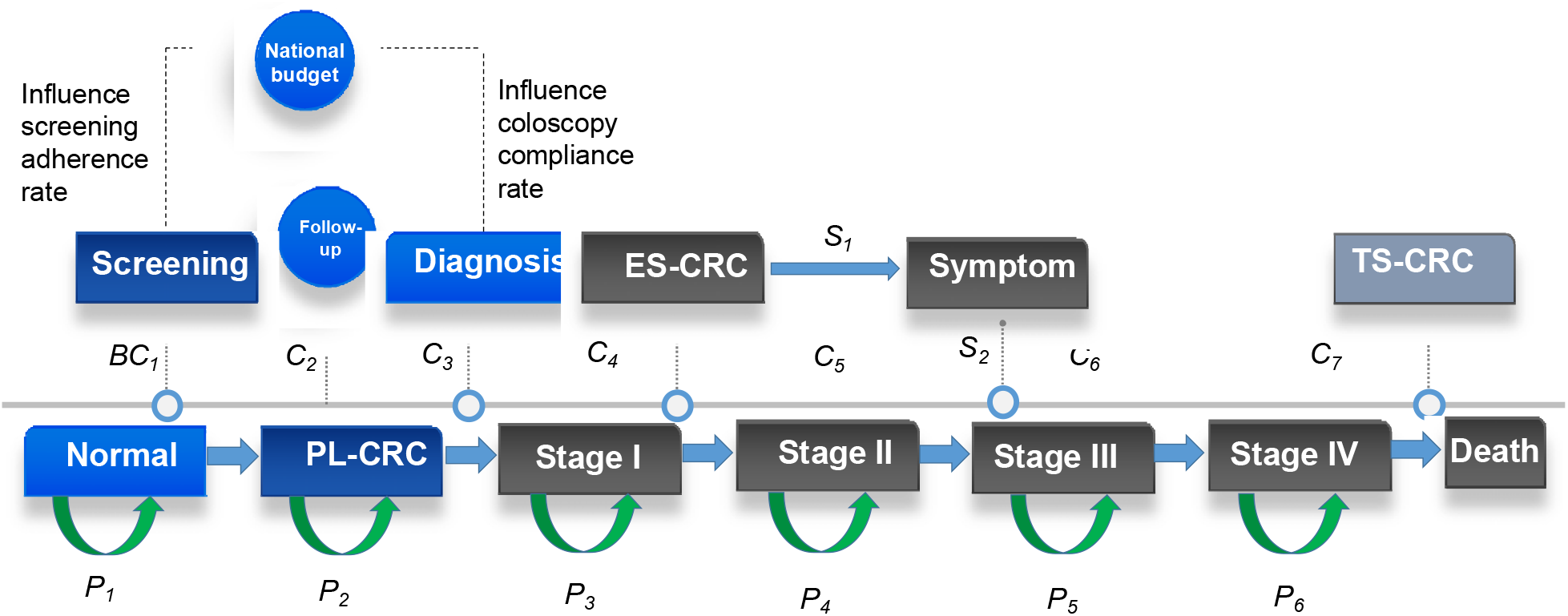
Flowchart of Colorectal Cancer Screening for Cost-effectiveness analysis. Legend: ES-CRC = Early stage colorectal cancer, TS-CRC = Terminal stage colorectal cancer, PL-CRC = Precancerous lesion colorectal cancer, *P*_*1*_ = Probability; *C*_*1*_ = *Cost 1*; *BC*_*1*_ = *BGI Cost 1; S1 = Symptom proportion 1*

If the screening fees was covered by government, the screening adherence was 5.8% for Fecal DNA methylation test and 6.6% for FIT. If the coloscopy was afforded by government, the compliance rate was 47% for Fecal DNA methylation test and 20% for FIT.

For the screening fees, Fecal DNA methylation test was charged 102.3 (*C*_*1*_) dollars as the direct cost for one person without labor cost, and FIT was 27 (*C*_*2*_) dollars. The early stage charges the cheaper treatment fees as 24,829 (*C*_*3*_) dollars because they might not present symptoms, and they could survive 4.5 years (*S*_*1*_) in this group. However, the treatment cost increases substantially to 326,237 (*C*_*4*_) dollars for the terminal stage after showing the clear symptoms, and the mean survival duration were 2.8 (*S*_*2*_) years in this stage.

### Optimal participation adherence

Under the assumption of optimal 100% participation adherence, Fecal DNA methylation test yielded the similar CRC incidence and the larger reduction in CRC mortality comparing than FIT (Table 2). Although CRC incidence was similar, the Fecal DNA methylation test-Positive cases were substantially higher than Fecal DNA methylation test-Negative cases for Stage I & II (35 vs 5), Stage III & IV (31 vs 4) and CRC cases (66 vs 9). FIT screening also shared the similar pattern with Fecal DNA methylation test.

**Table 2.** Base-Case Clinical and Economic Outcomes for Hypothetical 100,000-Person Cohorts with Perfect Screening Participation from Age 45 to 75 in Saudi Arabia

|  | Fecal DNA methylation test |  |  | FIT |  |  |
| --- | --- | --- | --- | --- | --- | --- |
|  | Positive | Negative | Total | Positive | Negative | Total |
| Status, number of cases per 100,000 persons (% of all cases) |  |  |  |  |  |  |
| PL-CRC | 2300 | 2700 | 5000 | 1200 | 3800 | 5000 |
| Stage I & II | 35 | 5 | 40 | 28 | 12 | 40 |
| Stage III & IV | 31 | 4 | 35 | 27 | 8 | 35 |
| CRC cases | 66 | 9 | 74 | 55 | 20 | 75 |
| CRC deaths per 100,000 persons |  |  |  |  |  |  |
| Total | 66 | 18 | 84 | 55 | 40 | 95 |
| Total cost (Million) | 34.52 | 884.16 | 918.68 | 22.04 | 1247.17 | 1269.22 |
| QALYs/person | 6.18 |  |  | 6.40 |  |  |
| Cost/person | 345.16 | 8841.59 | 9186.75 | 220.45 | 12471.71 | 12692.16 |
| Incremental cost/QALYs | 1487.30 |  |  | 1982.42 |  |  |

**Table 3.** the Predicated Number of Cases for Hypothetical 100,000-Person Cohorts with the Organized Screening Participation from Age 45 to 75 in Saudi Arabia

|  | FDMT |  |  | FIT |  |  | Ratio |
| --- | --- | --- | --- | --- | --- | --- | --- |
|  | Positive | Negative | Total | Positive | Negative | Total | FIT/<br>FDMT |
| Status, number of cases per 100,000 persons (% of all cases) |  |  |  |  |  |  |  |
| Total |  |  |  |  |  |  |  |
| CRC cases in 3 years | 189 (76.83%) | 57 (23.17%) | 246 | 1646 (87.93%) | 226 (12.07%) | 1872 | 7.61 |
| CRC cases in 6 years | 422 (85.77%) | 70 (14.23%) | 492 | 3514 (93.86%) | 230 (6.14%) | 3744 | 7.61 |
| CRC cases in 9 years | 665 (90.11%) | 73 (9.89%) | 738 | 5386 (95.90%) | 230 (4.10%) | 5616 | 7.61 |
| Early stages |  |  |  |  |  |  |  |
| CRC cases in 3 years | 176 (79.28%) | 46 (20.72%) | 222 | 1509 (87.63%) | 213 (12.37%) | 1722 | 7.76 |
| CRC cases in 6 years | 394 (87.56%) | 56 (12.44%) | 450 | 3222 (93.72%) | 216 (6.28%) | 3438 | 7.64 |
| CRC cases in 9 years | 620 (91.45%) | 58 (8.55%) | 678 | 4939 (95.81%) | 216 (4.19%) | 5155 | 7.60 |
| Terminal-stages |  |  |  |  |  |  |  |
| CRC cases in 3 years | 13 (54.17%) | 11 (45.83%) | 24 | 137 (91.33%) | 13 (8.67%) | 150 | 6.25 |
| CRC cases in 6 years | 28 (66.67%) | 14 (33.33%) | 42 | 292 (95.42%) | 14 (4.58%) | 306 | 7.29 |
| CRC cases in 9 years | 45 (75.00%) | 15 (25.00%) | 60 | 447 (96.96%) | 14 (3.04%) | 461 | 7.68 |
| CRC deaths per 100,000 persons in total |  |  |  |  |  |  |  |
| CRC deaths in 3 years | 10 (40.00%) | 15 (60.00%) | 25 | 58 (45.31%) | 70 (54.69%) | 128 | 5.12 |
| CRC deaths in 6 years | 36 (58.06%) | 26 (41.94%) | 62 | 236 (69.62%) | 103 (30.38%) | 339 | 5.47 |
| CRC deaths in 9 years | 96 (76.19%) | 30 (23.81%) | 126 | 1391 (90.56%) | 145 (9.44%) | 1536 | 12.19 |

Yearly Fecal DNA methylation test yielded the lower mean QALYs of 6.18 per person than FIT every year. Comparing with FIT screening, Fecal DNA methylation test every year was less costly (918.68 million vs 1269.22 million), whereas Fecal DNA methylation test saves the average of $9186.75 for each person through home and society, and FIT charged $12692.16. In ICER comparisons between two strategies, Fecal DNA methylation test was more effective as 1487.30 per QALY gained than that of FIT every year 1982.42 per QALY gained (Table 2).

### Organized participation adherence

We assumed the organized Fecal DNA methylation test program triennually, 5.8% screening adherence (5800 cases) and 47% compliance rate (2726 cases) for colonoscopy after Fecal DNA methylation test-Positive in Figure 2. For the organized FIT screening program, it was annual screening for 6.6% screening adherence (6600 cases) and 20% compliance rate (1345 cases) after FIT-Positive based on the previous Saudi Arabia screening program. The final four outcomes had normal, precancerous lesions, early stages colorectal cancer and terminal stages colorectal cancer.

**Figure 2.**
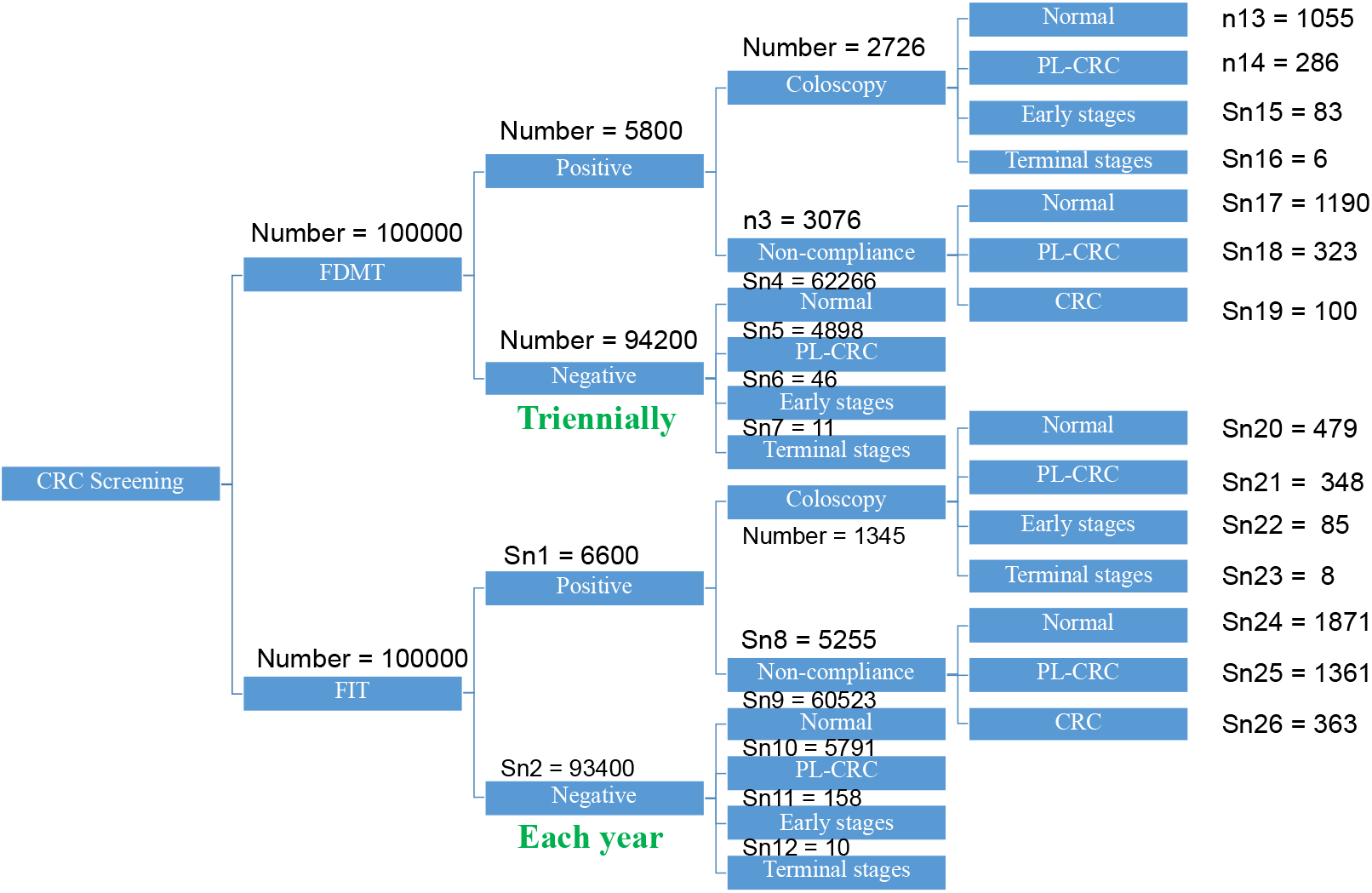
Compare of Fecal DNA methylation test and FIT Screening. **Legend:** Snx = Simulated number of status, which x denotes the set (1, 2, … 26)

For the Sn compartment, it meant the simulated number of cases for each status with the total of 26 compartments in Figure 2. According to the predicted outcomes, the normal status accounted for the largest proportion among four outcomes from 479 cases to 62266 cases. The terminal stages colorectal cancer had the smallest proportion from 6 cases to 11 cases. Relatively, the early stages colorectal cancer had 4.2 times to 15.8 times than terminal stages. Furthermore, the FDMT had the lower number of CRC cases than FIT screening both for early stage (89 cases vs 93 cases) and terminal stages (57 cases vs 168 cases) under the assumptions of FDMT.

### Predicated Number of Cases

In cohorts of population following by each screening cycle, our model simulated the predicated number of 5507 CRC precancerous lesions for the organized Fecal DNA methylation test screening program and 7500 CRC precancerous lesions for the organized FIT screening program in the first year. Furthermore, we also simulated the total number of CRC cases for Fecal DNA methylation test within three years (246), six years (492) and nine years (738). However, the FIT screening program was sharply increased to 1872 cases for three years, 3744 cases for six years and 5616 cases for nine years. Finally, we simulated the number of deaths between Fecal DNA methylation test and FIT within three years (25 vs 128), six years (62 vs 339) and nine years (126 vs 1536). Furthermore, the early stage had the larger number of cases than the terminal stages (Relative risk range: 6.25-7.76).

## Discussion

This study firstly clarified the cost effectiveness research of the novel fecal DNA methylation test (FDMT) comparing with the common FIT test. Our research proved the potential tool of the original FDMT test that is more sensitive than FIT to screen for colorectal cancer. The results interpret that at the perfect and organized screening adherence, FDMT is likely less costly and more effective than FIT. Since our prediction model interprets that FDMT could reduce the larger CRC incidence and mortality rates than FIT, depending on triennial and annual screening adherence, it is more likely to be selected, because FDMT might be the acceptable alternatives of FIT in Saudi Arabia.

In Saudi Arabia, the compliance of colonoscopy is extremely low as 9%, because it was invasive and more expensive than FIT or FDMT, even though colonoscopy is the golden standard to confirm Colorectal cancer. To reduce the Colorectal cancer burden in Saudi Arabia, we recommend the parallel tests including colonoscopy, FIT and FDMT to establish the more reasonable screening system in Saudi Arabia. In Saudi Arabia, the national budget might influence the willingness for both population screening adherence and coloscopy compliance.

To reduce the burden of colorectal cancer and improve the compliance of screening, the FDMT sensitivity and specificity were firstly estimated in Qingdao city-level screening program. In the local health department, they recruited 18136 participants for FDMT test to screen colorectal cancer in Qingdao. This program initially identified 1050 positive cases as 5.8% (1050/18136) initial positive rates for FDMT, and 493 of which had colonoscopy adherence rate as 47.0% (493/1050).

Overall, FDMT is the novel screening toolkit potentially beneficial to serve the larger population. Under the pressure of medical insurance in countries, this toolkit might be one of the valuable screening approach for the global market.

### Conclusions

In summary, two simulation results show that FDMT is likely to dominate FIT for colorectal cancer screening of the national program in Saudi Arabia. FDMT may be an alternative cost-effective tool for both perfect adherence and the organized participation, available to reduce the substantial burden of CRC in the Saudi Arabia population.

## Data Availability

All data produced in the present study are available upon reasonable request to the authors

## Declarations

### Consent for publication

Not applicable.

### Availability of data and materials

Not applicable.

### Competing interests

We declare that the research was conducted in the absence of any commercial or financial relationships that could be construed as potential conflicts of interest.

### Funding

Not applicable

### Contributors

Zhongzhou Yang and Shida Zhu designed the project and carried out this study. Zhongzhou Yang and Mengping Liu analyzed the data and prepared the manuscript. Eric Lau and Shida Zhu helped to draft the manuscript and provided the substantial suggestions to improve the manuscript. In addition, Mang Shi, Zhe Wang, Xiaoyuan Zheng and Yanyan Liu revised the manuscript together. Yantao Li and Zhongzhou Yang collected the parameters.

## Acknowledgements

We acknowledge BGI Saudi Arabia team like Jianjun Li and HKU team Zonglin Dai.

## Notes

### Competing Interest Statement

The authors have declared no competing interest.

### Funding Statement

This study did not receive any funding

